# Archaeological biological mineralizations: A cutting-edge multi-proxy investigation on kidney stones from the Argaric Bronze Age civilization

**DOI:** 10.1101/2025.11.26.25341105

**Authors:** Robin Francotte, Camila Oliart, Jennifer Bodin, Stéphan Rouzière, Cristina Rihuete-Herrada, Eva Celdrán Beltrán, Vicente Lull, Brecht Laforce, Jacob I. Griffith, Vince Laszlo, Christophe Snoeck, Steven Goderis, Michel Daudon, Emmanuel Le Tavernier, Dominique Bazin, Rafael Micó, Frederik Tielens

## Abstract

Two calculi retrieved at the archaeological site of La Almoloya (Murcia, Spain) dating to the Early Bronze Age were analyzed to evaluate their preliminary identification as kidney stones and to explore their pathological and social implications for the health status of a prehistoric complex society. The samples were examined through Scanning Electron Microscopy, X-Ray Fluorescence, X-Ray Diffraction, Fourier-Transform Infrared Spectroscopy and Micro Computed Tomography. Chemical results were combined with genetic and osteoarchaeological analysis. The recovered kidney stones are composed of primarily carbapatite, whitlockite and amorphous carbonated calcium phosphate. In one case, bacterial imprints were observed. Their chemical composition indicates an infectious origin; for one individual, the stone was probably formed due to a urinary tract infection, which was likely caused by a fracture of the ischiopubic ramus, which in turn can lead to urethral injuries. As urinary tract infections had persistent disabling effects on carrying out daily activities, the individuals involved would have benefited from social care. The study highlights the value of multidisciplinary approaches for identifying unusual bioarchaeological remains. Recovery of kidney stones in archaeological contexts remains challenging due to their small size and sedimentary resemblance; systematic screening of burial sediments could improve detection. DNA analysis of genetic material from pathogens preserved in kidney stones could help improve differential diagnosis and lead to a better assessment of the severity and persistence of the infection that caused the formation of the *calculi*.

## 1. Introduction

The population of El Argar, which inhabited southeastern Iberia during the Early Bronze Age (2200-1550 cal. BCE), provides one of the best scenarios to study the emergence of substantial population expansion and technological advancements during late prehistoric Europe [1–5]. The Argaric culture left behind a rich archaeological legacy characterized by large, permanent settlement, distinctive artefacts, advanced metallurgy [6], intensive farming and animal husbandry practices, an extensive funerary record (thousands of single and double graves located under the settled areas), and significant economic and political inequalities [7–11]. Researchers have used demographic and paleopathological analyses of human skeletal remains to investigate health conditions in Argaric times [12, 13].

Recent excavations in one of the paramount El Argar sites, La Almoloya (Region of Murcia, Spain), led to the discovery of several small biomineralized items of human origin in two graves [14]. The discovery in Argaric sites of calcified concretions, visually identified as urolithiasis according to their morphological traits, provides a new window into the health status of this prehistoric population. This type of *calculi* can result from a variety of factors, including dietary habits, anatomical singularities, metabolic disorders (e.g., systemic acidosis, low urinary volume, genetic predisposition to absorptive abnormalities), and urinary tract infections [15, 16]. Therefore, multidisciplinary scientific research is needed to reach a reliable differential diagnosis. In this study, we use Scanning Electron Microscopy (SEM), X-Ray Fluorescence (XRF), X-Ray Diffraction (XRD), Fourier-Transform Infrared (FTIR) and Computed Tomography (CT) to characterize the composition, morphology, and, hence, the origin of the biomineralizations yielded by two prehistoric individuals. These results were then combined with insights from archaeology, human osteology, palaeopathology, and medical science, aiming to reconstruct critical aspects of diet, lifestyle, medical knowledge, and health status of the Argaric 4000 years ago.

## 2. Materials and methods

### 2.1. Retrieving calculi from archaeological contexts

One of the main issues hindering a broader interest in pathological biomineralizations produced by past individuals is the difficulty to retrieve these items in archaeological contexts. The external features of the *calculi* make it difficult to distinguish them from the surrounding matrix, particularly in deposits with varied sedimentary textures and gravel-rich compositions [17]. Thus, to ensure the correct identification of such mineral deposits in the field, the excavation of funerary contexts should ideally be carefully conducted by personnel trained in osteoarchaeology. The location of the *calculi* in relation to the skeletal remains, particularly when preserved in articulation, is also important for the initial stages of differential diagnosis. In any case, it is strongly recommended not to discard after the excavation the sediments filling the grave, but to keep them for subsequent dry sieving [18] or, preferably, for water processing with a flotation machine because of the fragility of many calcifications, which requires careful procedures for retrieval, handling, and storage. To monitor diagenetic processes and contamination, a blank soil sample should also be collected before sieving.

The application of this protocol led to the retrieval of four *calculi* from two tombs at La Almoloya, discovered during the 2015 excavation campaign, within the framework of the Almoloya-Bastida Project, led by a research team from the Universitat Autònoma de Barcelona (ASOME-UAB). The team initially identified several small items as possible kidney stones, thanks to the careful digging and sieving of the sediment that filled tombs AY-60 and AY-68. Both were double graves in which an adult male and, afterwards, a female was buried. Radiocarbon dating shows that these four inhumations took place between ca. 1900 and 1800 cal BCE, during the second occupation phase of La Almoloya. Genetic analyses indicated that these individuals were biologically unrelated and found no biological markers of pathogens [19, 20]. Grave goods suggest that the individuals belonged to different social classes [21, 22].

In tombs AY-60 and AY-68, the skeletal remains of the first burial were removed to make room for the second body and were subsequently placed on top of it, covering its full length except for the head (Figure 1). This ritual procedure and the very fact that two corpses were placed in the same grave might *a priori* make it difficult to ascertain which individual the biomineralized items belong to. However, the tomb’s inner surface had been carefully cleaned, as shown by the absence of bones from the first body beneath the second. Along with the meticulous archaeological excavation, this observation enabled us to assign the two biomineralized items to the second individual in both graves.

**Figure 1.**
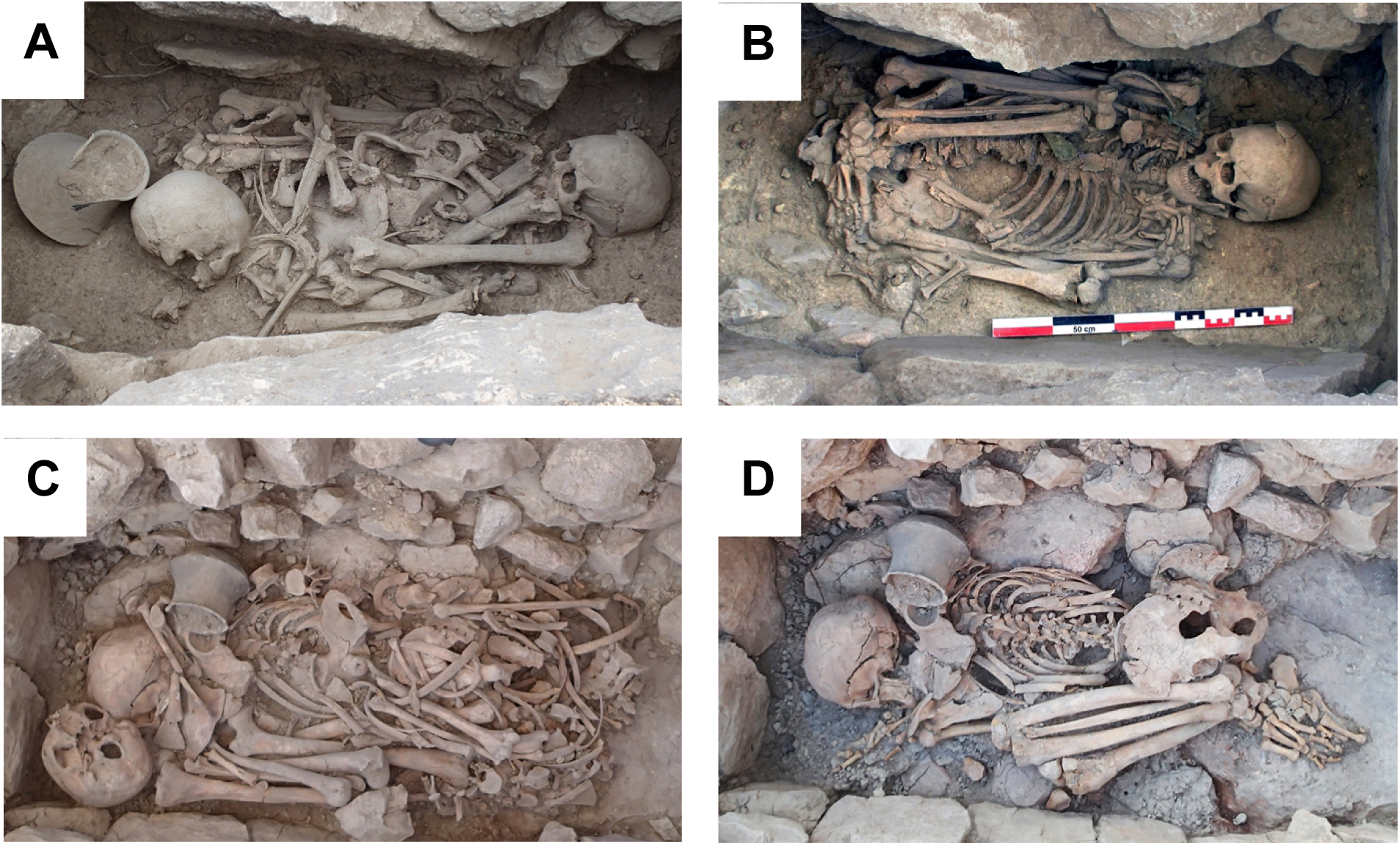
La Almoloya tombs AY-60 and AY-68. A: Commingled bones of the female individual AY-60/1 spread on top of the male AY-60/2. B: Male individual AY-60/2 in anatomical position). C: Commingled bones of the male individual AY-68/1 fully covering the remains of the female AY-68/2. D: Female individual AY-68/2 in anatomical position. Source: ASOME-UAB.

In tomb AY-60, the second burial (AY-60/2) was a mature/elderly male (>40 years of age-at-death based on the fusion of the jugular synchondrosis) [23] with several healed traumatic injuries to the coxal bone and skull that prevented the application of additional methods for age estimation. The individual was inhumed in a supine position with their legs extremely flexed and lying on either side of their body, in what appears to have been a very tight bundle. A *calculus* was recovered from the abdominal area. This item had an irregular shape, with a maximum length of 2.09 mm. The pale blush surface showed bulbous projections with its internal structure being fine-grained and ivory-white in color, resembling a “mulberry” or a “hemp seed”. The internal structure was fine-grained and ivory-white (Figure 2). Based on its morphology and location [24], it was initially identified as a renal stone. The sediment covering the skeletons was processed in a flotation machine to retrieve small elements and find fragmented remains that were undetected in the field excavation. As a result, two additional biomineralizations were found. Their shape and internal structure were similar, featuring also bulbous projections, but differed in size (8.72–6.46 mm and 14.22–5.35 mm) (Figure 2).

**Figure 2.**
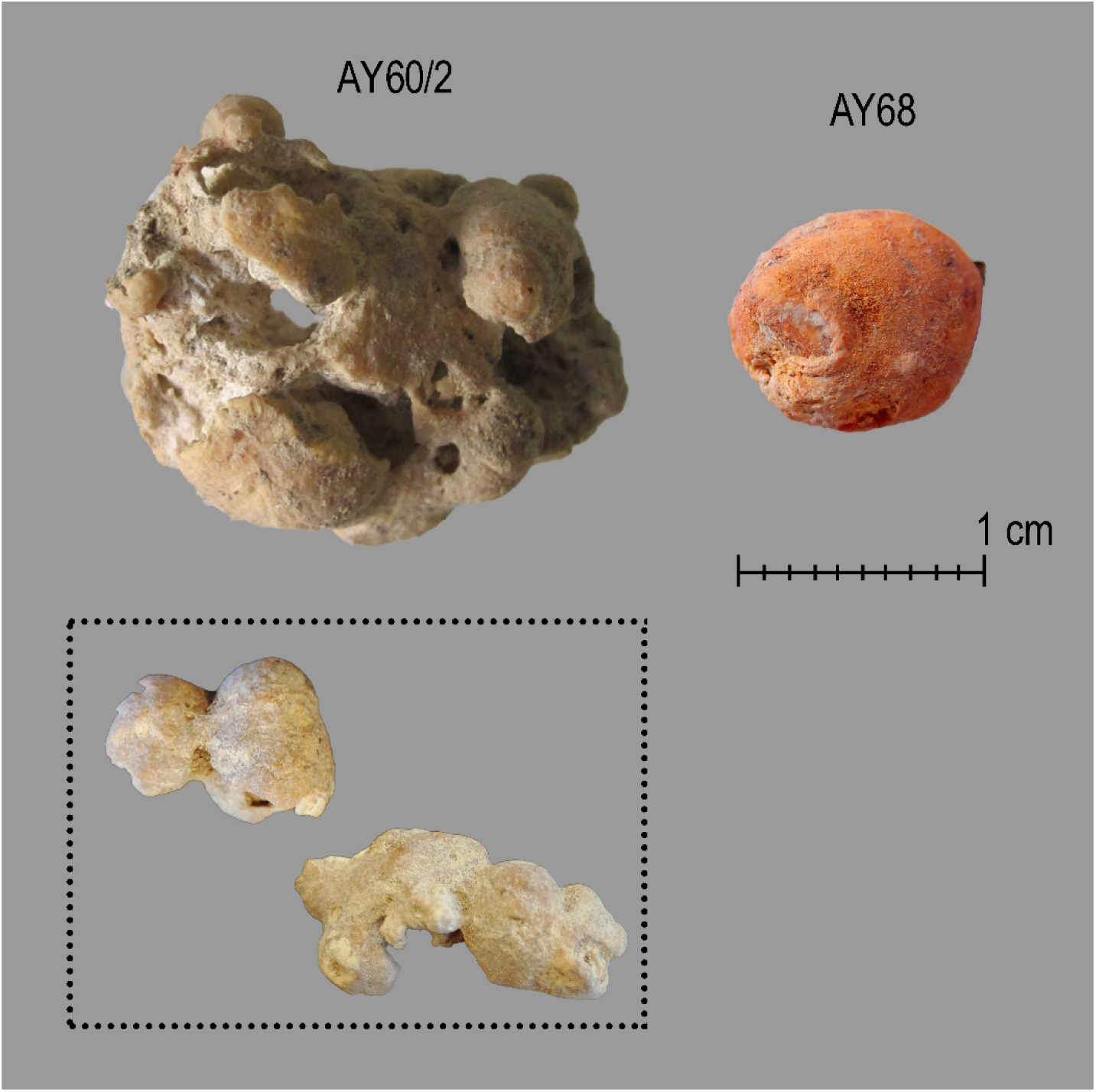
The large biomineralization found in association with AY-60/2 in the field (upper left); *calculi* retrieved after the flotation of the sediments filling the tomb AY-60 (inset, bottom left); calculus from AY-68 (upper right).

In tomb AY-68, the second burial was that of a biological female (AY-68/2), aged between 30-35 years based on the morphology of the pubic symphysis and the auricular surface [25–27], as well as the fusion of the sternal epiphyses of both clavicles and the complete fusion of the sacral vertebrae [28]. The skeleton was inhumed in a prone position with their legs tightly flexed and bent to the right. During the excavation of the pelvic and lower back areas, a nodule was found, measuring between 4.56 mm (minimum length) and 7.79 mm (maximum length). It had a smooth blush-white surface and a fine-grained, ivory-white internal structure (Figure 2). Based on its location and morphological features [24], it was identified as a urinary calculus.

The *calculus* recovered in the field from AY-60 provided three samples. Fragments AY-60/2-S1 and AY-60/2-S2 come from the core of the original *calculus*, while fragment AY-60/2 represents a large portion of the cortex. The material from AY-68 consisted of a single biomineralized item. In addition, soil samples from burial AY-60 were collected and analyzed to assess possible contamination of the *calculi* by its sedimentary environment.

### 2.2. A Study of Biomineralization

Initially, a set of chemical analyses was needed to test the nature of the possible biomineralizations and, subsequently, to offer the first clues of the processes leading to the formation of these items.

#### a. SEM observations

We employed SEM to investigate the microstructural characteristics of the kidney stones. This technique provides preliminary insights into the constituents of the stone by analyzing the morphology of surface crystals [29]. SEM is particularly valuable because it reveals details of the inner structure, crystal morphology, and possible bacterial imprints of urinary stones, thereby providing clues to the causes for their formation. The sample analyzed through SEM was AY-60/2-S2, which represents the inner portion of the item.

A Zeiss SUPRA55-VP scanning electron microscope equipped with an energy-dispersive X-ray (EDX) spectrometer has been used for direct microstructure observation of a sample without any conductive coating. This field emission gun microscope, which can operate at 0.5–30 kV accelerating voltage, allows for high resolution observations obtained at 1 kV using an Everhart–Thornley detector [30].

#### b. X-ray fluorescence experiments

Subsequently, (bulk) XRF was employed to determine the **elemental composition** of the samples, which is useful for elemental analysis and characterization of **trace elements** that could potentially be nephrotoxic (Cd, Pb …), and could therefore indicate, for instance, whether the individual lived near a mining site [31, 32]. Both calculi samples (AY-60/2, AY-60/2-S1, AY-60/2-S2, AY-68) were studied by XRF; the surrounding soil from burial AY-60 was also analyzed as a blank.

X-ray fluorescence (XRF) measurements were carried out on a setup of the MORPHEUS platform at Laboratoire de Physique des Solides (LPS - Université Paris Saclay/CNRS), comprising a microfocus molybdenum X-ray tube coupled with a multilayer mirror optics (Incoatec, GmbH), which delivers a monochromatic X-ray with energy equal to 17.48 keV, and a SDD (Silicon Drift Detector, Ketek GmbH) energy dispersive detector [33].

#### c. Micro-X-ray fluorescence analysis

Micro-XRF or µXRF enables the assessment of the **elemental distribution** across the sample surface and facilitates the identification of localized regions (hotspots) with increased concentrations of specific elements [34]. The samples studied by µXRF were AY-60/2-S1 and AY-60/2-S2.

Elemental distributions with (3D) micro-XRF in the HERAKLES 3D microprobe of Ghent University [35–37]. This multimodal equipment combines micro-Computed Tomography (µCT) with 3D elemental sensitive techniques to obtain integrated data sets revealing the internal elemental structure of microscopic samples. Experiments on HERAKLES went through three steps: (1) µCT mapping the internal structure of the samples; (2) high-resolution (20-30 µm) overview mappings using XRF yielding 2D mappings of the elemental distribution of all detectable elements; and (3) local 3D XRF analysis (XRF-CT) to identify the depth of discovered hot spots of elements of interest, determining whether they are located on the surface or deeper within the sample.

#### d. Micro-Computed Tomography measurements

To further examine the internal microstructure, µCT was conducted, allowing for the visualization of distinct phases within the sample [29]. Unlike SEM, µCT provides information on the distribution of main components through the stone and allows for the mapping of calcium phosphate, calcium oxalate and organic components from a whole stone. Only the sample AY-68 was studied with µCT.

μCT measurements were conducted on a Bruker 1172 Skyscan μCT (voxel size: 0.5 μm) housed in the Archaeological, Environmental Changes and Geochemistry Research Unit of the Vrije Universiteit Brussel. Due to issues with size and fragility, only AY-68 could comfortably fit into the scanning chamber and be scanned. The total scan time was set to 3 hours and 42 minutes. Source voltage and current were set to 100kVP/100µA, combined with a 0.5 mm aluminum filter and a 0.5 mm copper filter. The number of rows and columns were set to 2664 and 4000 respectively, producing 322 TIFF image files. The image pixel size was 6.73 μm, with a 0.6° rotation step, 360° tomographic rotation, and flat field correction enabled, with the type of motion set to ‘step and shoot’. The completed scan was rendered using CTAn v.1.18 software.

#### e. X-ray Diffraction experiments

Additionally, X-ray Diffraction (XRD) analysis was performed to determine the **mineralogical composition** of the sample [29, 34]. The obtained diffraction patterns were compared with reference data to identify the constituent mineralogical phases. We applied the method to both calculi (samples AY-60/2, AY-60/2-S1, AY-60/2-S2; AY-68) and the surrounding sediment from burial AY-60.

X-ray Diffraction (XRD) analyses were carried out on the MORPHEUS platform at the Laboratoire de Physique des Solides (LPS), with a home-made diffraction setup installed on a rotating anode X-ray generator (model MicroMax007, Rigaku) at the Cu wavelength (λCuKα = 0.1542 nm) delivered by a multilayer W/Si mirror (Xenocs). Diffraction patterns were collected on a large-area detector MAR345 (marXperts GmBH) with 150 µm pixel size placed at a distance of 250 mm from the sample. Extraction of diffracted intensity I as a function of the scattering angle was obtained from the azimuthal angular integration of the diffraction patterns with a home-made developed software [33].

#### f. FTIR analysis

FTIR was used to characterize the vibrational modes of the molecular components within the samples. The resulting spectra were analysed in conjunction with reference spectra of pure compounds, facilitating both qualitative and quantitative evaluation of the sample’s composition [38]. FTIR allows for the **identification of all components** in a sample, whether they are crystalline or amorphous, mineral or organic, provided they are present in sufficient quantities (0.5-1%). In addition, FTIR can be used to determine the **carbonation rate** of apatite in the stone, providing further insight into the disease that may have affected the individual. The inner portion of the calculus (AY-60/2-S2) was analyzed by FTIR.

Stone analysis was performed at Tenon Hospital following the methodology described by Daudon and Bazin in 2016 [39]. The sample was analyzed by Fourier transform infrared spectroscopy (Vector 22 infrared spectrophotometer, Bruker Optics, Champs-sur-Marne, France) in absorbance mode by accumulation of 32 spectra between 4000 and 400 cm^-1^, with a 4 cm^-1^ resolution. For this sample, the inner and surface compositions were established. The compounds were identified by comparison to reference spectra [40].

After chemical analyses confirmed the initial classification of the items as biomineralizations and, namely kidney stones (*infra*), we applied additional approaches to delve further into their formation processes. The paleopathological study of skeletons AY-60/2 and AY-68/2 was conducted through osteoarchaeological and genetic analyses. Genetic data related to both individuals were available from a previous study [19] that found no specific pathogens. We have further explored a potential genetic basis in the formation of those kidney stones. Specifically, we focused on the variants of certain single nucleotide polymorphisms (SNPs) of gene CLDN14, rs219781[C], rs219780[C], rs219779[C], rs219778[T], linked to calcium reabsorption [41, 42]; and gene CaSR, rs181725[T] and rs1801197[G], that leads to decreased tubular calcium reabsorption within the nephron, resulting in enhanced urinary calcium excretion [42–44].

## 3. Results and Discussion

### 3.1. Urolithiasis

#### SEM observations

Crystal morphology of calculi depends on internal and external factors such as the size, shape and orientation of the seed [45]. For kidney stones, using the link between morphology and the chemical phase is challenging because of the large number of phases identified in them. Also note that the same chemical phase may lead to different kinds of morphology. Such is the case for calcium oxalate monohydrate, which displays a crystal morphology depending on the pathology that induced the kidney stones [46].

The different multiscale observations performed through SEM indicate that the surface of the calculus AY-60/2-S2 is comprised of spherical entities which are probably made of calcium phosphate apatite, as well as some bacterial imprints as indicated by the red arrows in Figure 3a [47]. Some crystallites with a pseudo cubic morphology can also be observed (blue arrows, Figure 3b), which can be related to whitlockite [48].

**Figure 3.**
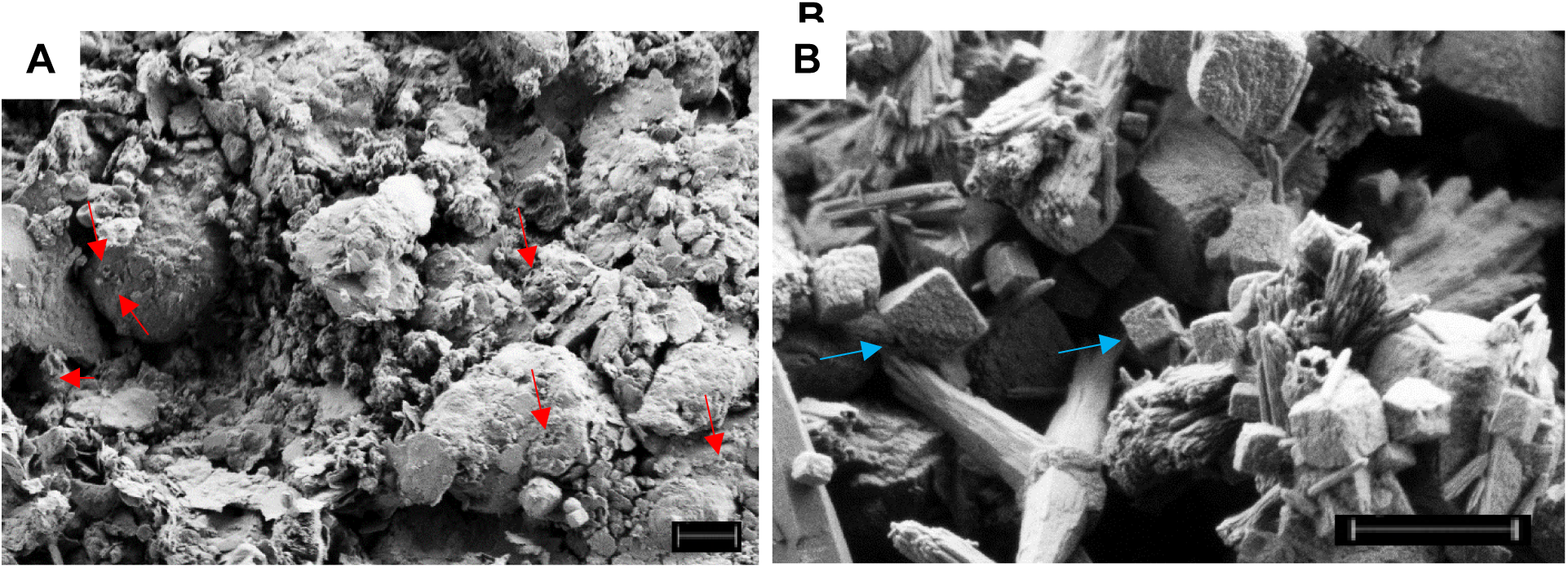
SEM images of sample AY-60/2 with a magnification of (A) 4000 X (scale bar = 2 µm) and (B) 20000 X (scale bar = 1 µm).

The presence of whitlockite in the kidney stone sample suggests that the stone was likely formed due to an infection, such as a urinary tract infection (UTI), especially if it is detected alongside carbapatite or struvite [48]. The bacterial imprints clearly hint at a bacterial infection, which again could be related to a UTI in the presence of carbapatite [49]. It is, however, important to mention that not all apatite stones are necessarily related to an infectious origin [50].

#### XRF measurements

The elementary composition can be derived from the different X-ray fluorescence spectra, which clearly show the presence of calcium and phosphorus, indicating that these calculi are primarily composed of calcium phosphate (Figure 4 and Table 1) [51]. Other elements such as Zn, Cu, Fe, Sr, Rb and Se are also present in kidney stones [31]. In this case, elemental peaks for Ti, Cr, Mn, As, and Br have also been identified. µXRF analysis confirmed the presence of several of these elements.

**Figure 4.**
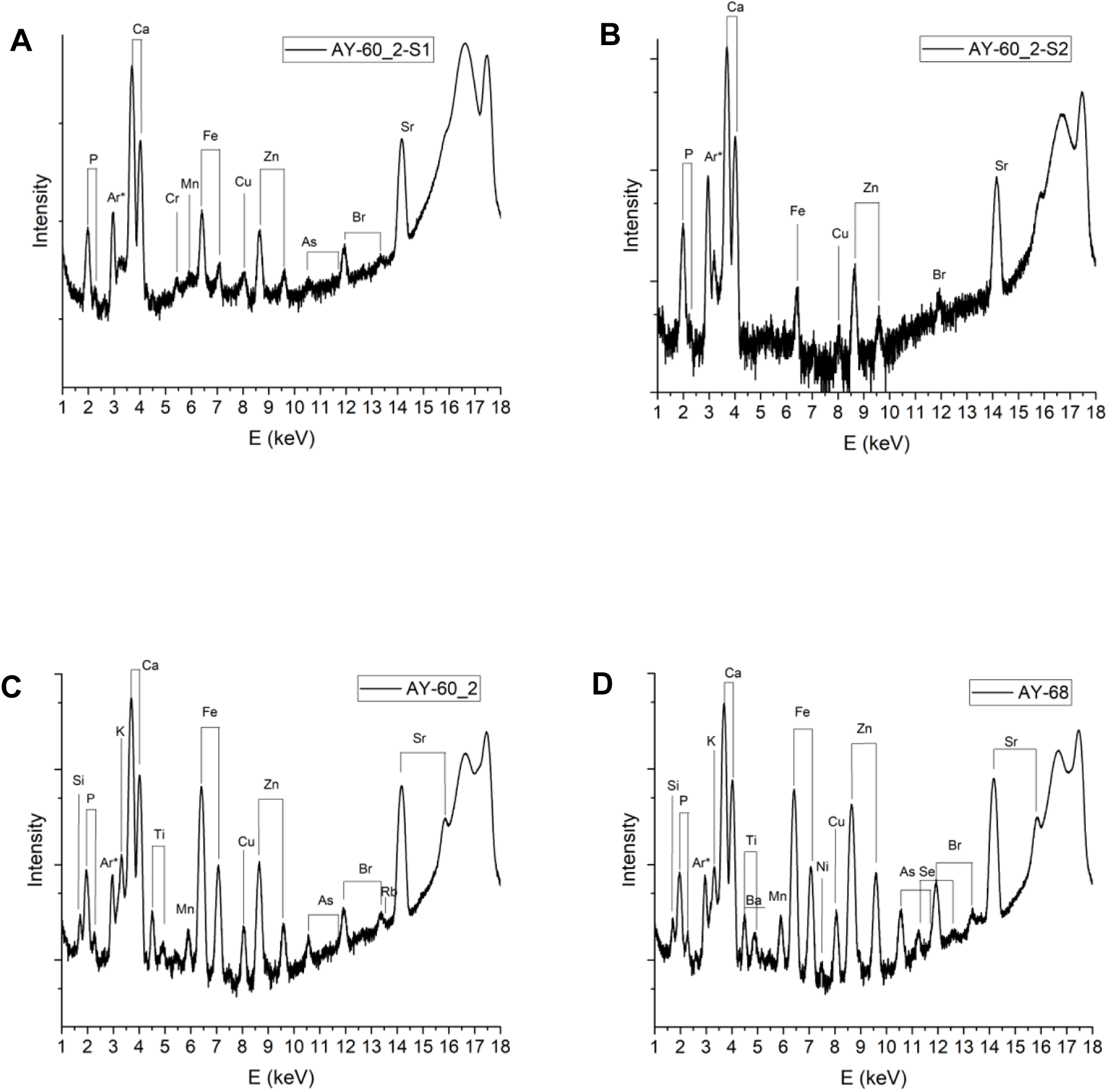
Bulk XRF spectra of AY-60/2-S1 (A), AY-60/2-S2 (B), AY-60/2 (C) and AY-68 (D).

**Table 1.**
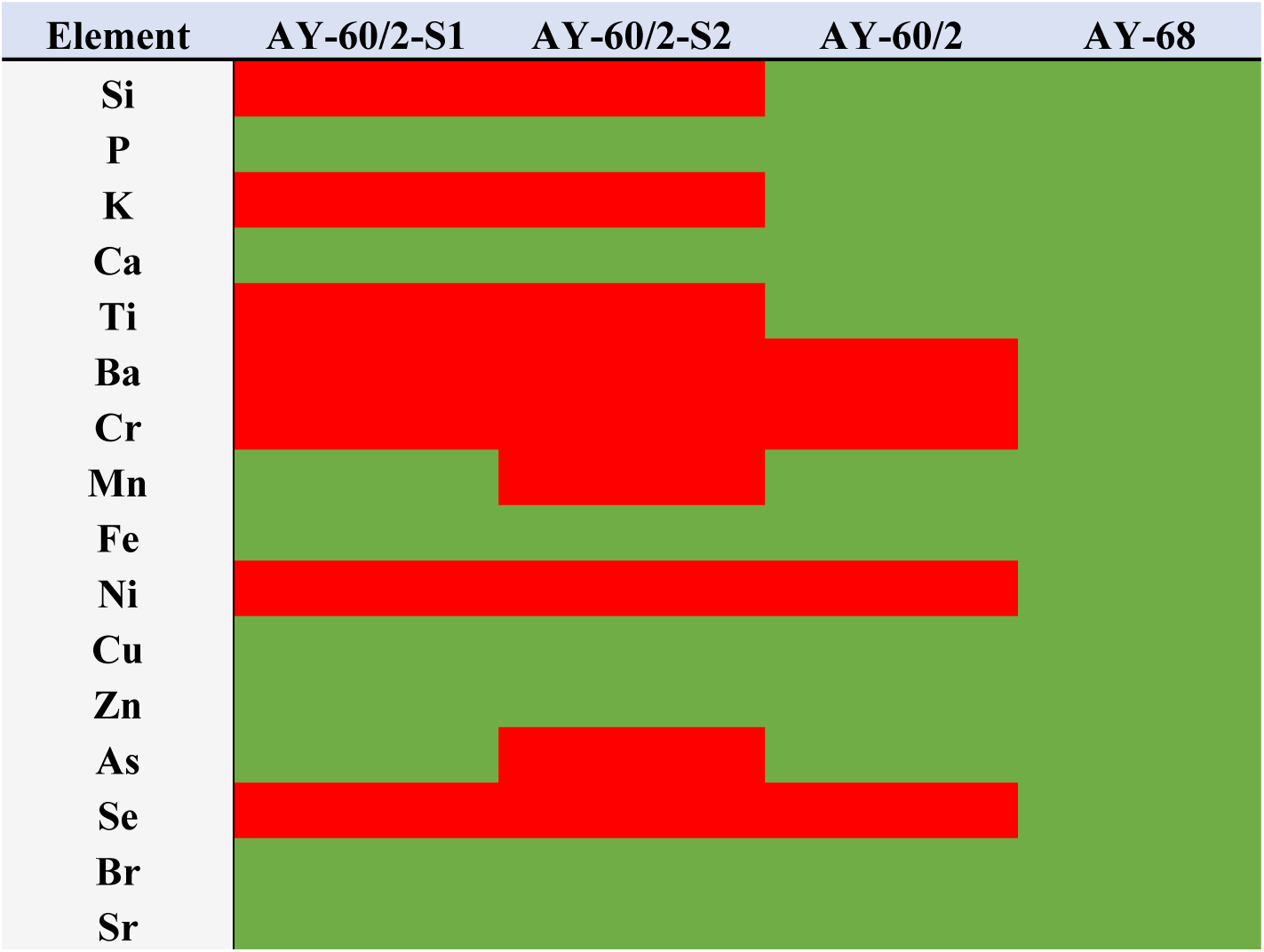
Overview of the elements detected in the calculi using XRF. Green color means the element was detected using bulk XRF, while red color means that the element was not detected.

Different investigations have been performed regarding the precise role of trace elements in the pathogenesis of kidney stones [52]. For example, trace elements may modify the speed of the crystallization process and influence the external morphology of growing crystals [53]. An investigation on 78 calculi of well-defined composition by means of µXRF analysis using synchrotron radiation as a probe, allows us to assess the presence of trace elements in kidney stones [31]. As an example, the presence of Zn and Sr in calcium stones in the crystallographic network is likely to result from the similarity in ion charge and radius with calcium, favoring a substitution process of calcium by metal ions rather than a contribution to stone formation. As Zn and Sr are more abundant in calcium phosphate than in calcium oxalate stones, these elements also suggest a high abundance of calcium phosphate in the studied samples. Fe, Cu and Rb are typically much less abundant, but have also been found in calcium stones. Other elements such as Ti, Mn, and As may come from the soil environment and their presence is thus likely the result of diagenetic processes.

Based on the XRF spectrum of the soil in burial AY-60 (Figure 5), the elemental composition of the soil differs from that of the kidney stone samples obtained from burial AY-60 as most notably P was not detected in the soil.

**Figure 5.**
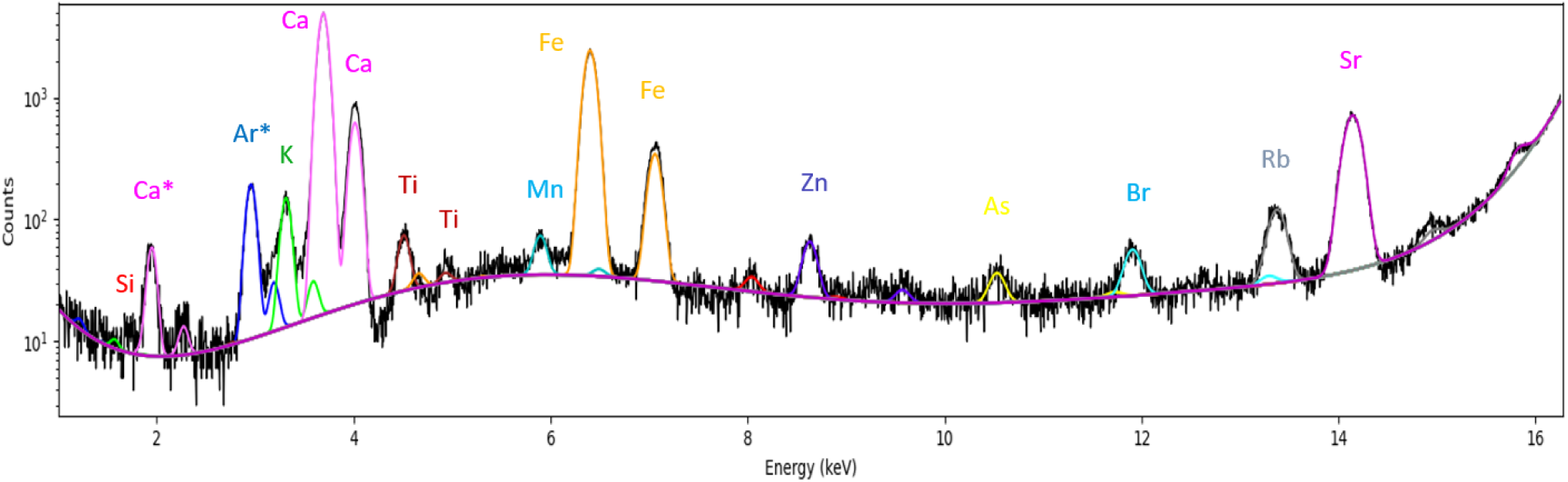
Bulk XRF spectrum of soil collected from burial AY-60.

#### 3D µXRF analysis

3D µXRF analyses were performed with the aim of tracing the distribution of the different elements across the biomineralized items. In this way, elements coming from contamination of the calculi (by the sediments that filled the tombs, for example) instead of from the actual individual can be distinguished. Following respective sizes, only samples AY-60/2-S1 and AY-60/2-S2 were analyzed by 3D µXRF.

As can be seen in the overview scans, µXRF confirms the high concentrations of Ca and P. Secondarily, the presence of strong Fe, Sr, and Zn concentrations is affirmed, while smaller quantities of K, Ti, and Ni are observed (Figure 6). The increased signal near the edge of the items in these images is artificial edge enhancement. Interpreting the results is different for the lower atomic number elements such as P and the metals, since information depth increases with the mass of an element. In all cases the signal is clearly stronger towards the rim of the sample. Considering the edge effect, all slices indicate a relatively homogeneous distribution of Ca, P, Sr, and Zn, likely confirming a substitution of Ca, while this does not appear to be the case for K, Ti, and Fe (and possibly Ni), for which hot spots are observed in the individual XRF slices. These hot spots, with a maximum diameter of ~0.5 mm, may indicate the presence of adhering material (for example in the case of Ti in Figure 6a or because of diagenetic processes, as supported by the detection of Ti in the soil blank. Direct comparison to a set of modern kidney stones would be required to discriminate diagenetic effects from trace contributions of other mineral phases or metals to stone formation, since these samples were buried alongside bone tissue in soil.

**Figure 6.**
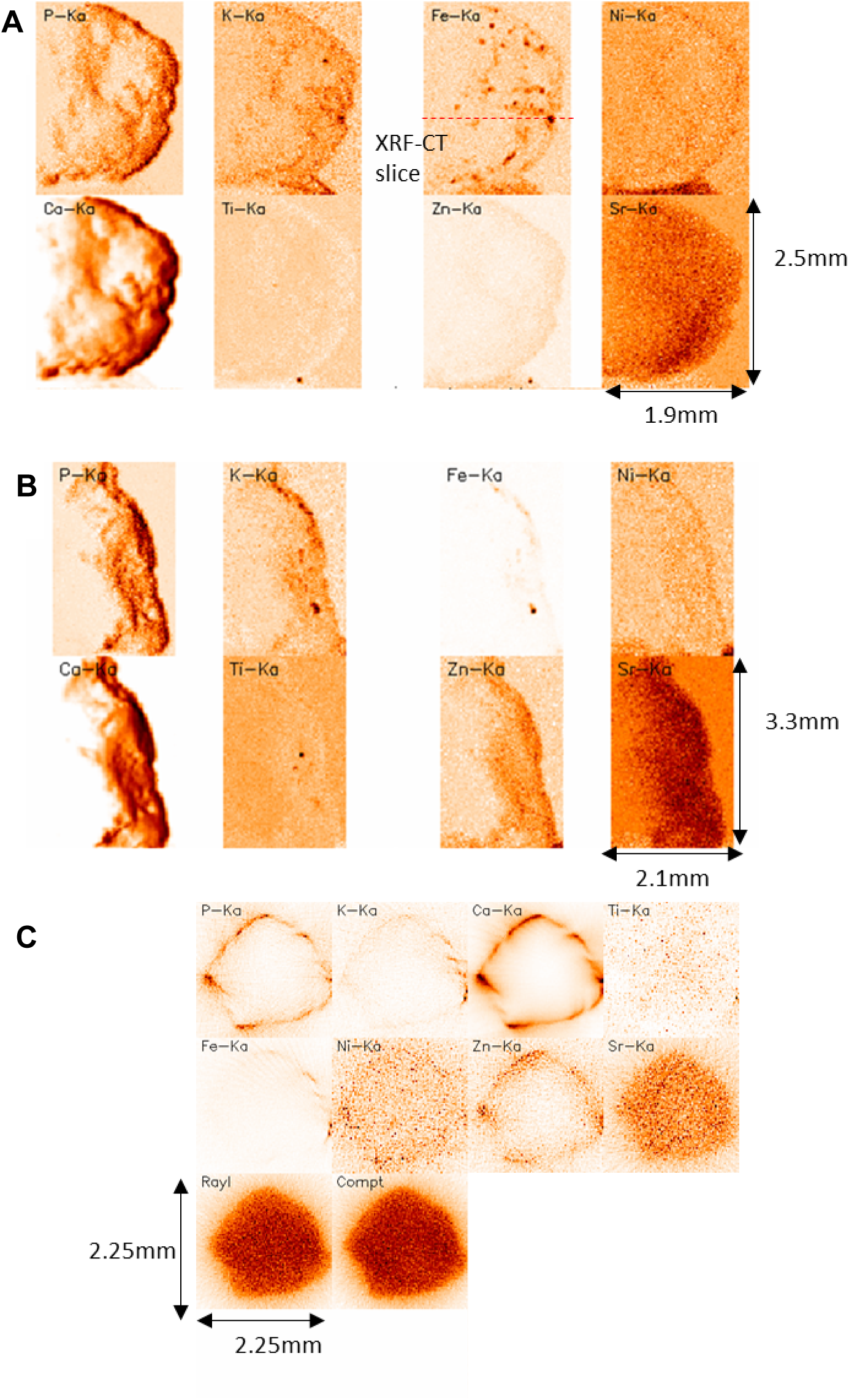
Results from the overview µXRF mapping of sample AY-60/2-S1 (A, 20 µm step size) and AY-60/2-S2 (B, 30 µm step size) and the XRF-CT slice through the sample at the position indicated (C, 25µm step size) for a variety of elements, including P, K, Fe, Ni, Ca, Ti, Zn and Sr. Darker colors mean a stronger signal and higher concentration.

#### µCT measurements

Micro computed tomographic imaging (µCT) remains relatively novel in the field of biomineralization analysis [54]. This characterization technique is, currently, not able to give information regarding the chemical phases present in kidney stones. This is because some chemical compounds, for example all the urates, 1-methyl uric acid, dihydroxyadenine as well as stone matrix and uric acid have a similar X-ray attenuation value. However, µCT can still provide an indication of which compounds might be present in the sample.

The μCT rendering of AY-68 reveals the structure of the stone to have uneven points of density. The outer layers of the stone are denser in their inorganic structure than in the earlier phases in the core. Interestingly, this density is lopsided, with one focal point to the edge (Highlighted Point A: Figure 7) that holds the greatest structural density. Thus, point A (Figure 7) could consist of a concentrated spot of carbapatite, as it is known to have a high structural density [55]. The presence of carbapatite indicates that the pH of the urine was elevated, which could be the result of a UTI (especially if the carbonation rate of the carbapatite is high) or metabolic disorder.

**Figure 7.**
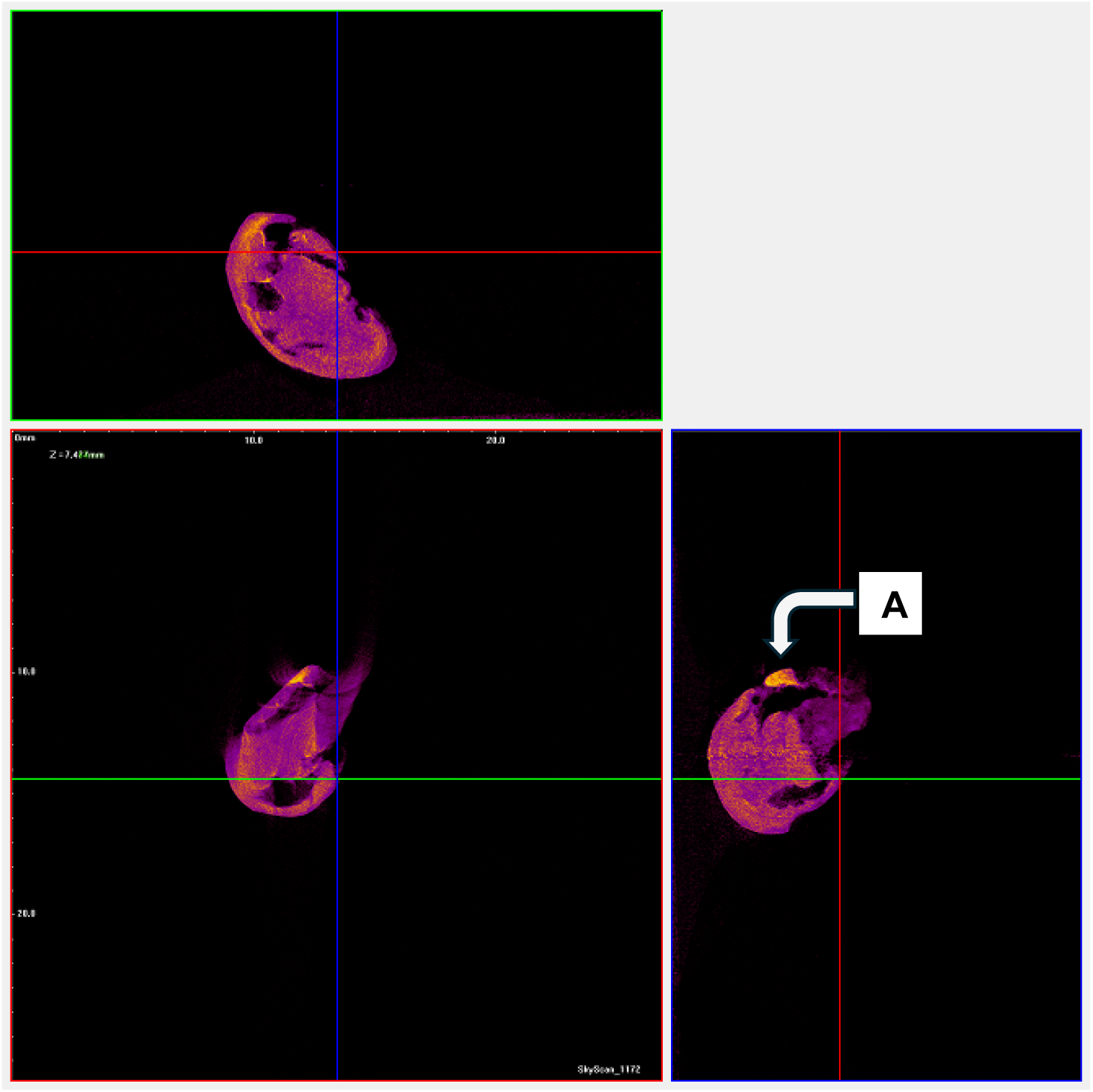
µCT images of sample AY-68. Brighter colors signify an area of higher density.

#### XRD measurements

Using XRD, a clearer understanding of the mineral phases present in the analyzed samples can be obtained.

In Figure 8, different diffractograms of the four samples are presented. In order to gain more understanding of the structure, the diffractogram of sample AY-60/2 is compared with diffractograms of hydroxyapatite (HAp) and whitlockite in Figure 9.

**Figure 8.**
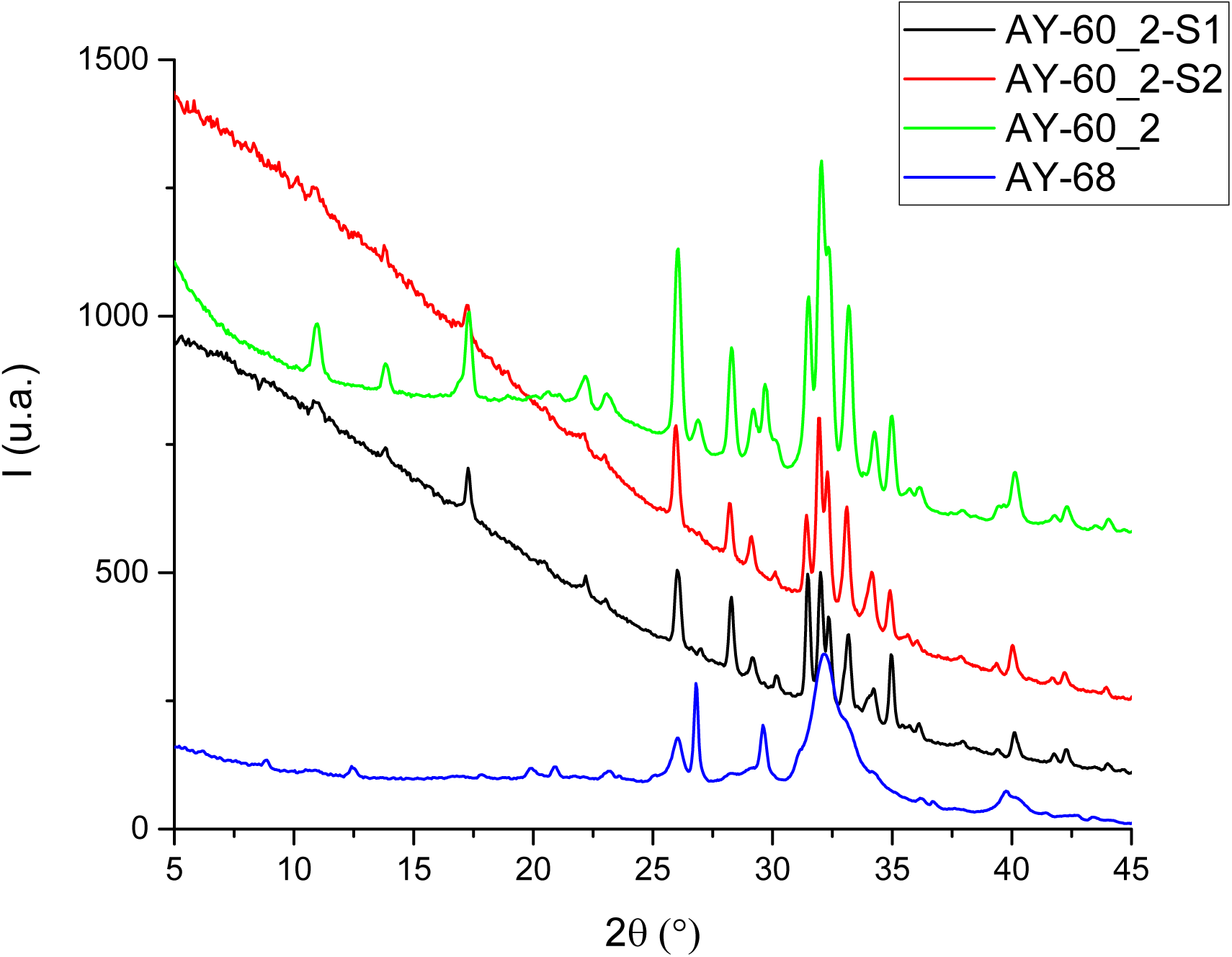
Diffractograms of AY-68 (blue), AY-60/2 (green), AY-60/2_S1 (black) and AY-60/2_S2 (red).

**Figure 9.**
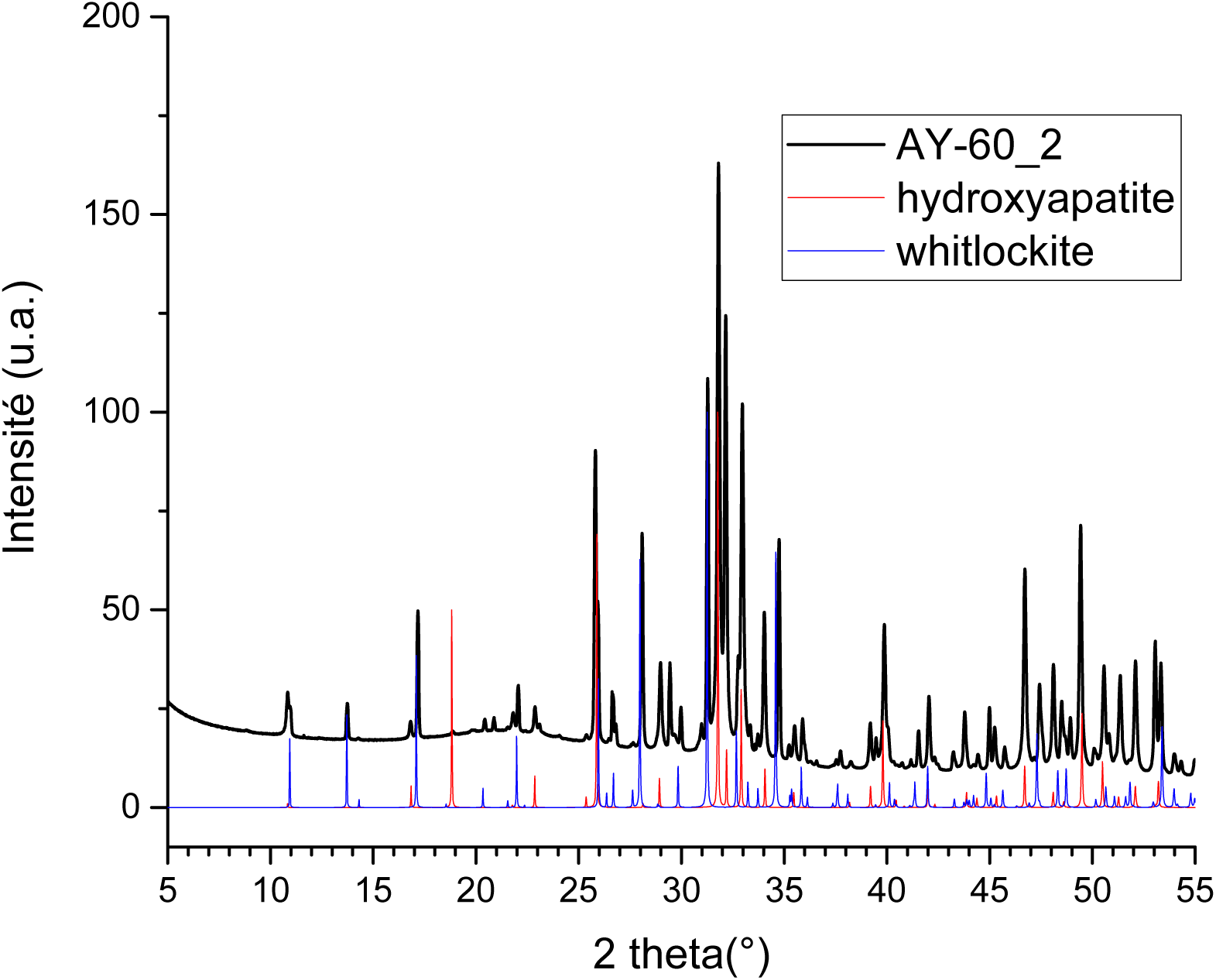
Comparison of the diffractograms of sample AY-60/2, hydroxyapatite and whitlockite.

From Figure 9 it is immediately apparent that the diffractogram of sample AY-60/2 is strikingly similar to that of hydroxyapatite and whitlockite, indeed suggesting the appearance of these two phases in the sample. This is not entirely unexpected, as both whitlockite [48] and hydroxyapatite [56] can be present in urinary stones, especially if the stone has an infectious origin, and was already hinted at through the SEM observations. HAp presence indicates that the urine pH is elevated, which could be the result of renal tubular acidosis, hyperparathyroidism or, alongside a high carbonate content, could indicate an infection such as UTI.

Figure 10 shows that the diffractogram of the soil samples from burial AY-60 is considerably different from that of sample AY-60/2, as expected based on the XRF spectrum in Figure 3. Figure 11 reveals the composition of the soil to be consisting mostly of calcite, dolomite and quartz.

**Figure 10.**
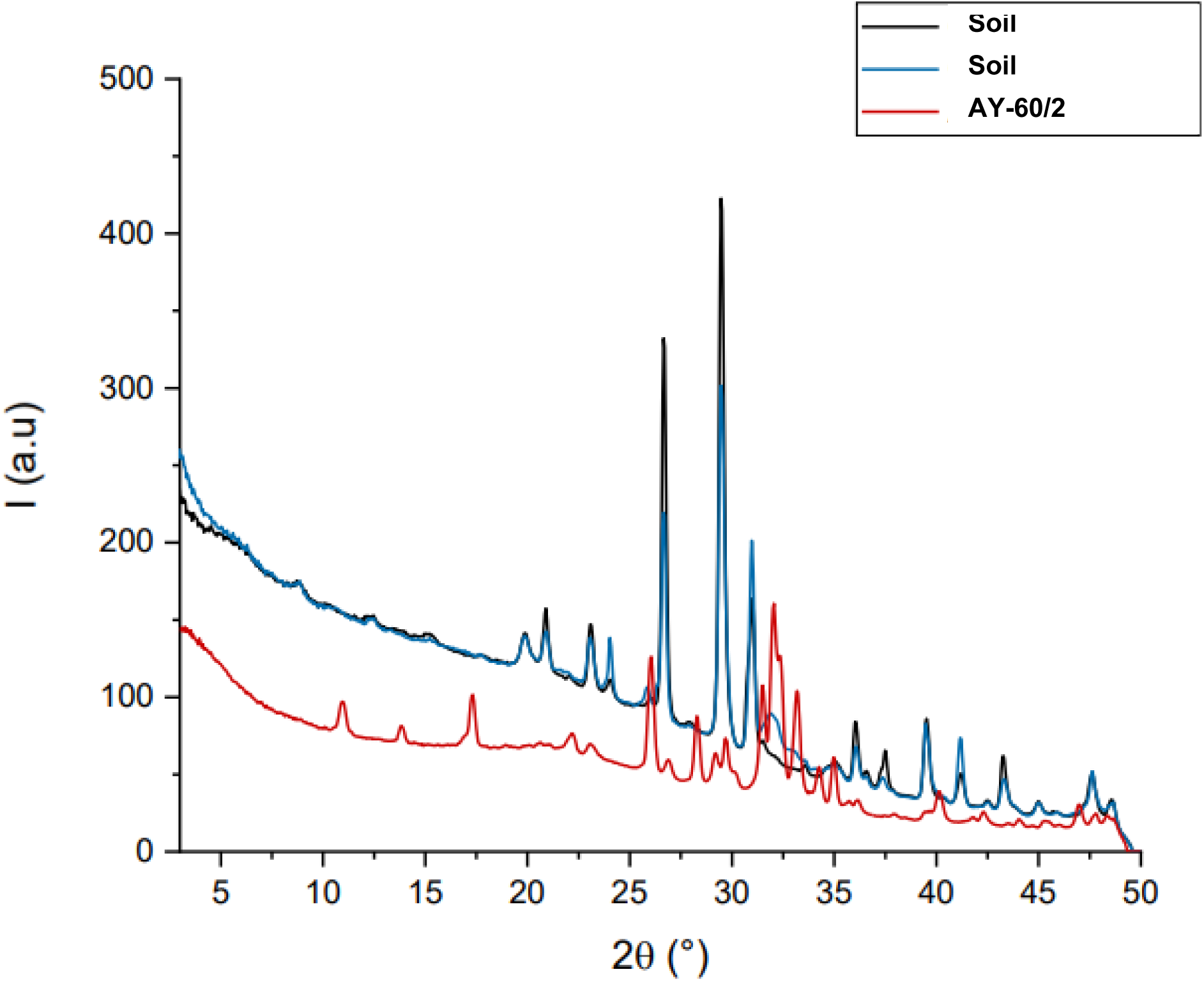
Comparison of the diffractograms of sample AY-60/2 (red) and surrounding soil samples (blue and black).

**Figure 11.**
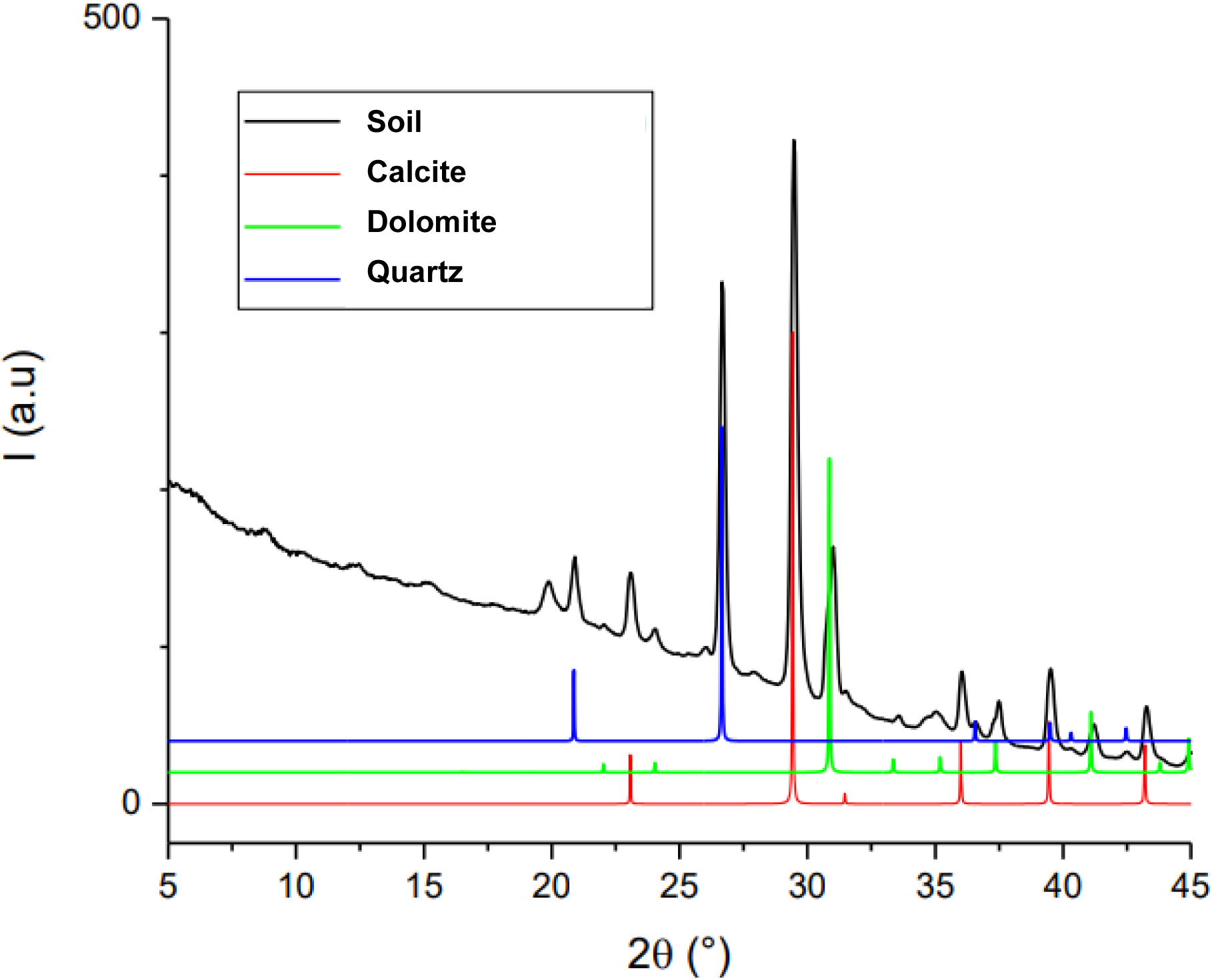
Comparison of the diffractogram of soil collected from burial AY-60 and the diffractograms of calcite, dolomite and quartz.

#### FTIR measurements

FTIR allows for both a qualitative and quantitative determination of the minerals present in the calculi, as well as the carbonation rate of apatite. For the FTIR analysis, the sample needs to be in the form of a powder. This means that the original sample will be (partly) damaged. For this reason, sample AY-68 was not analyzed here, as it was determined that the sample was in too good of a shape to damage it. As a result, only sample AY-60/2-S2 was analyzed by FTIR.

The results from the following experiments coming from FTIR are in line with those of the elementary composition determined by XRF and the structural data obtained by XRD. At the hospital, the morpho-constitutional model as given by M. Daudon establishes a link between the physicochemistry of kidney stones and the pathology which leads to their formation [57]. From the FTIR measurements (Figure 12), a global quantitative composition of the samples was determined, which is shown in Table 2.

**Figure 12.**
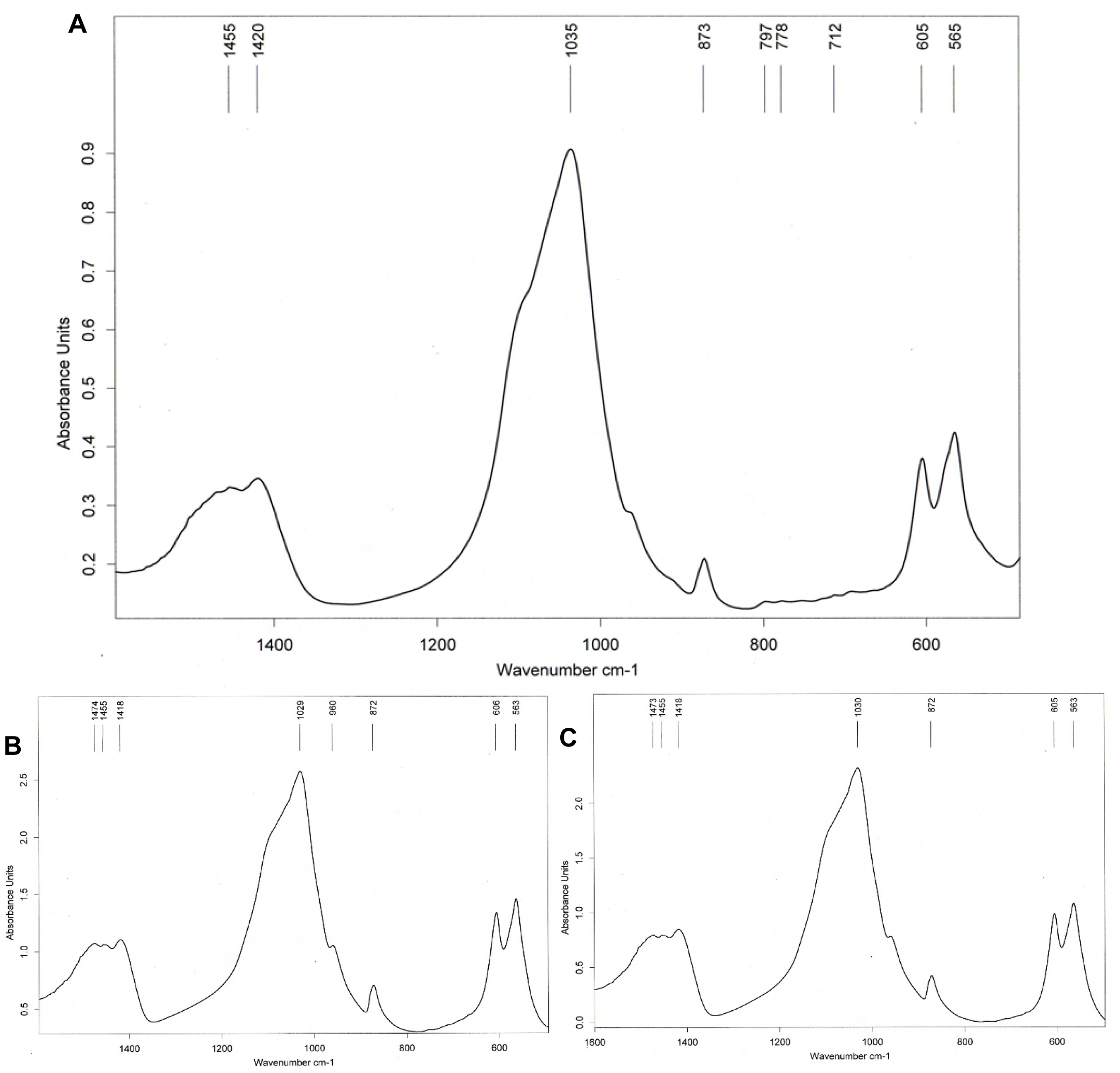
FT-IR spectrum of **A**) the surface layers, **B**) the middle layers and **C**) the deep layers of AY-60/2-S2, composed of calcium phosphate (carbapatite) mixed with whitlockite and amorphous carbonated calcium phosphate (ACCP). Since all layers show very similar signals, only figure A will be discussed here. Peaks at 1035, 605 and 565 cm^-1^ correspond to phosphate absorptions. Peaks at 1420–1455 and 873 cm^-1^ denote carbonate ions [38]. The shoulder of the peak at 1035 cm^-1^ indicates the presence of whitlockite. These results align well with the theoretically obtained values for phosphate, carbonate and whitlockite [58–60].

**Table 2.**
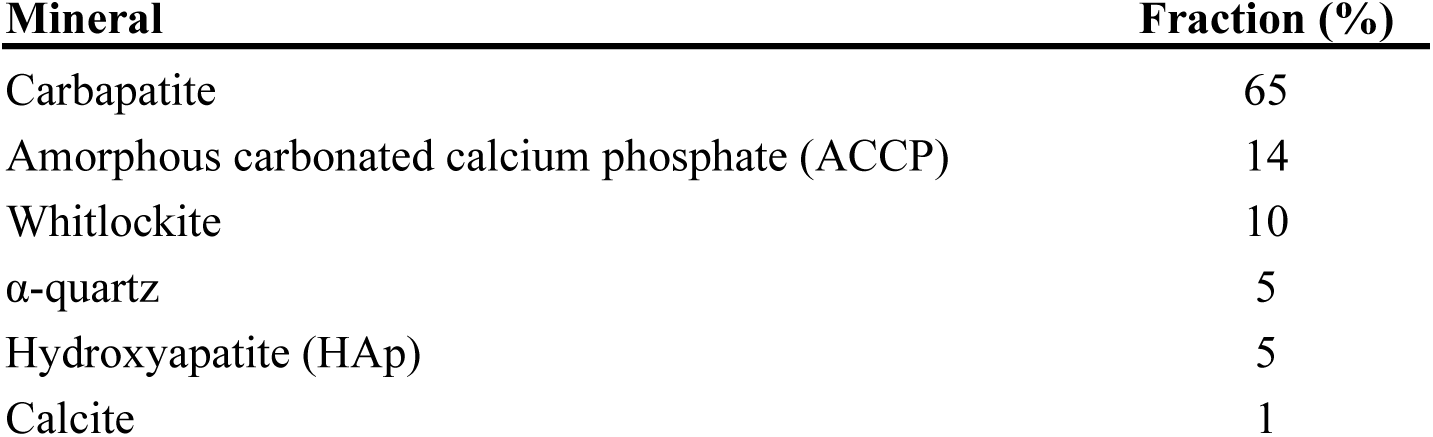
Global composition of AY-60/2-S2 as determined by FTIR.

| **Mineral** | **Fraction (%)** |
| --- | --- |
| Carbapatite | 65 |
| Amorphous carbonated calcium phosphate (ACCP) | 14 |
| Whitlockite | 10 |
| α-quartz | 5 |
| Hydroxyapatite (HAp) | 5 |
| Calcite | 1 |

Phosphate stones were highly prevalent among stone-formers in the past centuries and were often associated with urinary tract infection (UTI) and poor nutritional conditions both in adults and in children [61]. Calcium phosphate stones are characteristic of communities, such as those of the Neolithic period, where the diet was progressively richer in cereal carbohydrates and poorer in animal proteins [62].

Information regarding the disease comes from the carbonation levels of apatite [63]. For sample AY-60/2-S2 the carbonation rate was determined to be between 30% (in the surface layer) and 34% (in the inner layer). Such investigation indicates clearly that the finding of both a high carbonation rate and ACCP spherules in a phosphate stone, in the absence of struvite (which would have decomposed into trimagnesium phosphate pentahydrate or newberyite, both of which were not detected here), is highly suggestive of the past or current implication of microorganisms with weak urease activity.

Regarding our data, the chemical composition as given by FTIR spectroscopy leads to the fact that the stones are essentially composed of highly carbonated carbapatite, which suggests a chronic infectious origin (possibly caused by urea-splitting microorganisms) with a little ACCP, whitlockite and hydroxyapatite in the center and α-quartz on the surface (likely coming from the soil, see Figure 11). Should brushite or octacalcium phosphate have been detected alongside carbapatite, that could have been an indication of hypercalciuria, but neither of these components were found in our samples.

To summarize: the results from the chemical analyses obtained so far are consistent with the identification of the biomineralizations as kidney stones originating from an infectious process, especially in the *calculi* associated with AY-60/2. Concerning AY-60/2, SEM observations revealed spherical calcium phosphate apatite structures, pseudo-cubic whitlockite crystallites, and bacterial imprints, suggesting a possible infectious origin. Both XRF and µXRF analyses confirmed high concentrations of Ca and P, homogeneous distributions of Zn and Sr with some Fe, K and Ti hotspots. For AY-68, µCT imaging showed heterogeneous density, with denser outer layers possibly corresponding to carbapatite accumulation, indicative of elevated urinary pH. XRD also evidenced elevated urinary pH due the presence of HAp. Finally, FTIR spectroscopy corroborated these results, revealing high carbonation levels (30–34%) and ACCP spherules.

### 3.2. Genetic and osteoarchaeological findings

Variations in the CLDN14 and CaSR genes of the individuals AY-60/2 and AY-68/2 suggest low risk of kidney stone formation, with the only variant potentially influencing bone resorption being SNP rs219780, homozygous for the C allele (Table 3). Therefore, a genetic predisposition for the formation of urolithiasis can be ruled out.

**Table 3.** Summary of the SNPs analyzed in the CLDN14 and CaSR genes for the two individuals (AY-60/2 and AY-68/2).

|  | **CLDN14** |  |  |  | **CaSR** |  |
| --- | --- | --- | --- | --- | --- | --- |
| **IND/SNP** | **rs219778** | **rs219779** | **rs219780** | **rs219781** | **rs181725** | **rs1801197** |
| AY-60/2 | AA | GG | CC | GG | GG | AA |
| AY-68/2 | AA | GG | CC | GG | GG | AA |

The funerary goods in tomb AY-60 belong to a male of the Argaric ruling class: a copper halberd, a dagger, two portions of meat, and a ceramic vessel [14, 64]. The disposal of the two metal weapons, halberd on the right shoulder and dagger on the chest, just below the right knee, fits the pattern followed by the members of the warrior elite [65]. Not by chance, the male skeleton in AY-60 clearly reflects a long-term involvement in violent events. This includes two healed blunt force fractures on the right and left parietal bones, a traumatic erosion on the frontal bone, and a healed oblique fracture on the distal end of the left ulna that could be identified as a parry fracture [66]. The antemortem compression on the right lateral body of the axis and severe osteoarthritis in the lower cervical vertebrae may be associated with some of these traumatic injuries. However, the healed fractures in the vertebral portion of the body of two middle ribs (one right and one left), in the acromion of the left scapula, the intra-articular fracture of the distal articulation of the second phalanx and in the right coxae could have resulted from either an accident (i.e. after a fall from a moderate height) or interpersonal violence. There is no way to know whether these injuries were caused by a single or multiple events. Whatever be the case, this male benefited from intensive social care several times/for an extended period; moreover, the injuries led to some chronic physical limitations and probably pain. He died long after injuries had healed.

The trauma of the right hip is potentially associated with the formation of a urinary stone. The bone exhibits a healed fracture featuring extensive bone remodeling of the ischiopubic ramus and possibly the iliopubic ramus as well (Figure 13). There is no evidence that the right coxofemoral articulation nor the left hip were affected. Additionally, the poor preservation of the sacrum makes it difficult to determine whether the slight asymmetry of the posterior arch was caused by trauma. In any case, the injury did not affect the sacroiliac joint. This type of fracture can be classified as Later Compression type I (LC-I) according to the Young and Burgess system or, given that it affected a mature/elder male, as a Fragility Fracture of the Pelvis type 1a (FFP-1a) [67]. It was likely caused by low-force trauma, resulting in instability of the anterior pelvic ring. The callus and remodeling of the ischiopubic ramus have altered the morphology of the obturator foramen and correlate well with other physiological injuries, such as those affecting the pelvic floor muscles, the perineal membrane, the internal pudendal artery and/or the dorsal nerve of the penis. Moreover, bone spurs are present in the insertions of the sacrotuberous ligament, adductor magnus and obturator internus. Overall, it is likely that the urinary tract of AY-60/2 was damaged, posing a high risk of urinary infections [68], and even erectile dysfunction. Furthermore, the presence of no fewer than three renal *calculi*, measuring between 8 and 21 mm, likely led to episodes of renal colic, pyrexia, and possibly even loss of renal function.

**Figure 13.**
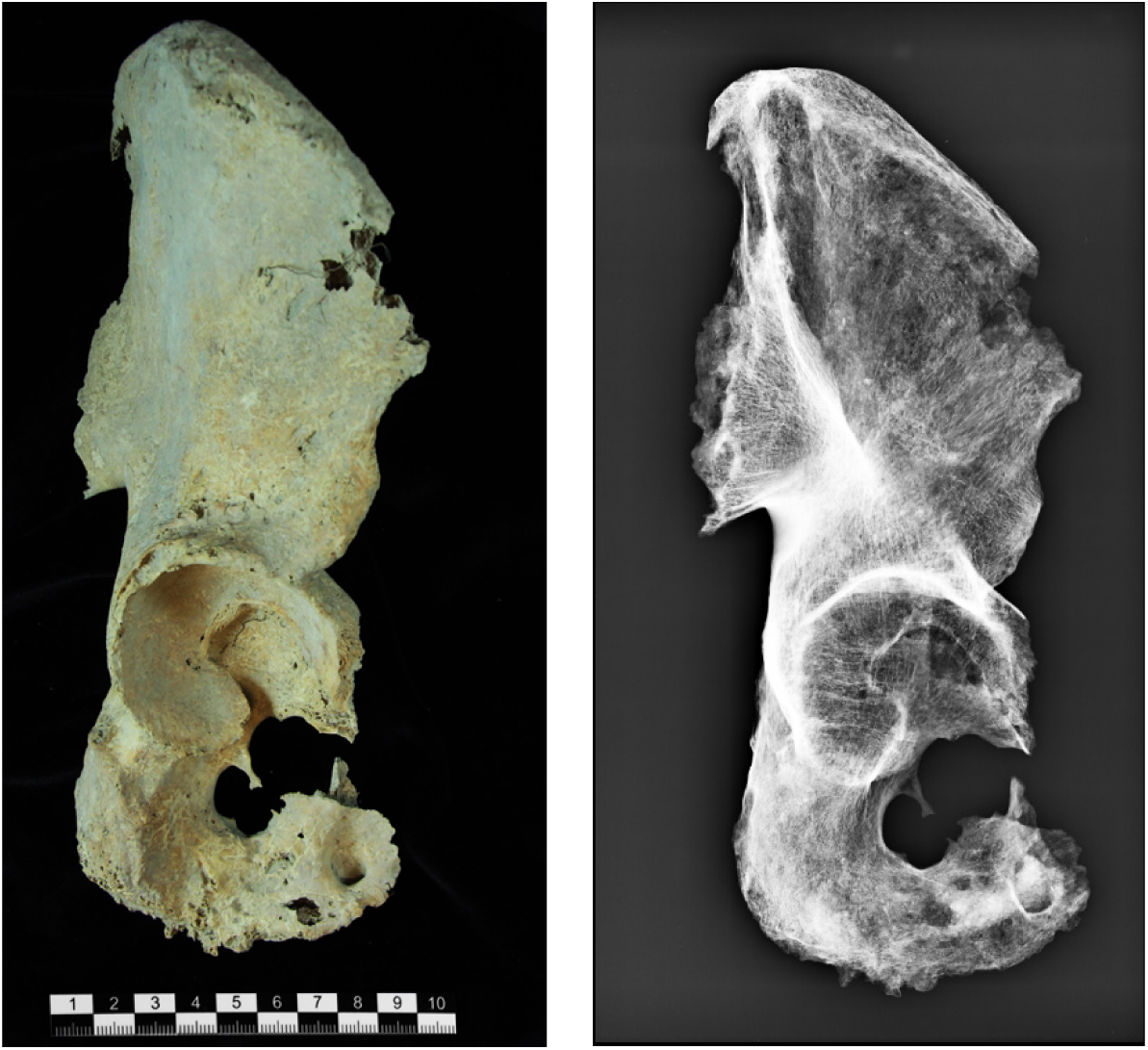
Photography (left) and radiographic image (right) of the right coxae with a consolidated fracture of the ischial ramus, obtained using a Toshiba Sedecal APR-VET unit (Model E7884X, maximum voltage: 150 kV). Radiographic parameters: vertical projection, 46 kVp, 12.5 mAs (400 mA, 0.032 s). Source: ASOME-UAB.

The grave goods found in tomb AY-68 were poor. Nevertheless, the presence of typical green stains from copper corrosion on the male’s right femur medial condyle and the sternal end of one rib, indicates that the first corpse had been in contact with at least one copper-based object. However, this object was not found among the bones of the first individual buried, which had become commingled and fully disarticulated due to the later disposal of the female. The only recorded funerary offerings were a pottery vessel and a portion of meat [64]. Although these objects do not provide clues about the tasks or social roles this individual performed during her lifetime, the archaeological study has been revealing. She died before reaching full biological maturity (around 30 to 35 years old). However, by the time of her death, she had already lost all her upper anterior teeth and most of the lower posterior ones, while the upper molars remained intact. This loss of upper anterior teeth is rather unusual, as the first molars are typically the first ones to be lost in adulthood. In addition, traces of extra-masticatory wear in the form of a groove on several mandibular teeth (central incisors, second incisor, and right first premolar) imply the use of the mouth as a ‘third hand’ in textile production (Figure 14), possibly during spinning yarn or any other activity involving fiber strands [9]. The use of teeth as a “third hand” (i.e., as a tool) may increase the risk of infections due to bacterial exposure, a close relationship between dental wear marks and the formation of dental calculi, reinforcing the role of bacterial activity. In this case, while the dental calculi are mild, the presence of periodontitis, caries, and antemortem tooth loss indicates poor oral health. She also suffered from other health issues, as shown by healed fractures in some ribs, a toe phalanx, and the first and second right metacarpals. The fractures in the metacarpals would have particularly hindered textile work, as they affect the fingers involved in pinching and gripping. Furthermore, she exhibits spondylolysis and a compression fracture of the fifth lumbar vertebra. Despite being a relatively young individual, other remarkable features include the fusion of the xiphoid process and the calcification of costal cartilages and the thyroid cartilage. In the case of AY-68, however, none of the osteological features observed can be unambiguously linked to health problems associated with the formation of a urinary stone.

**Figure 14.**
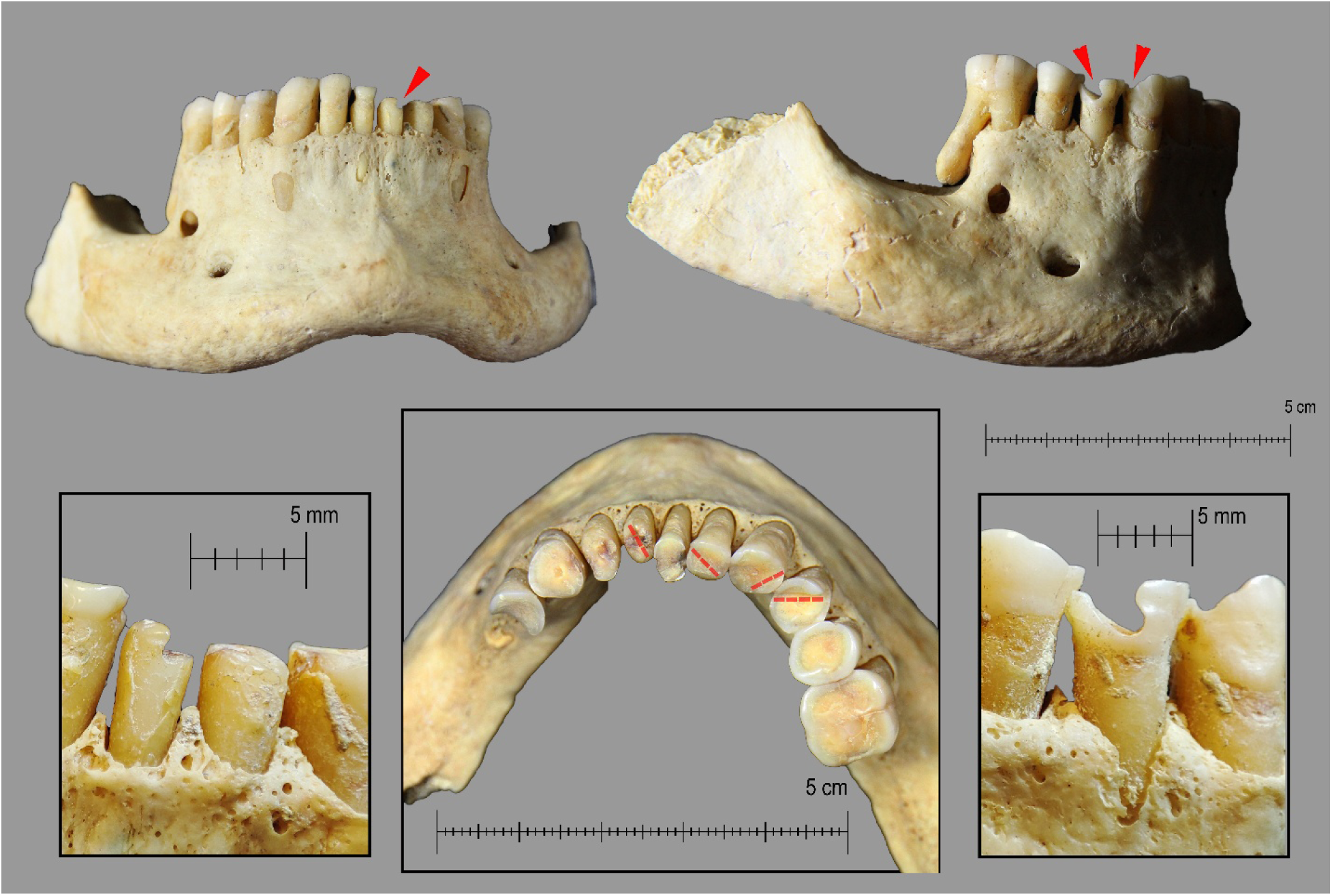
Extra-masticatory wear on the mandibular teeth of the female skeleton in AY-68. In the top panel, notches are indicated with arrows: the left central incisor shows a visible notch on its buccal surface (top left), as do the right first premolar and canine (top right). The bottom panel highlights the mandible with visible occlusal grooves. Source: ASOME-UAB

## 4. Conclusion

Multidisciplinary research has allowed for a more comprehensive approach to the living and working conditions of two Early Bronze Age individuals from the Argaric site of La Almoloya. The physicochemical characterization of two *calculi* recovered in the field confirmed to be kidney stones, as they were primarily composed of carbapatite, ACCP, and whitlockite. The complete set of data, gathered after SEM, XRF, µXRF, µCT, XRD, and FTIR analyses, indicates that these kidney stones had an infectious origin. Interestingly, in the absence of struvite, our data strongly suggests the presence of microorganisms with weak urease activity, while current day kidney stones of infectious origin are mostly formed due to microorganisms with high urease activity.

For the male buried in tomb AY-60, a clear connection between the results of the different analyses, including the osteological record, and the origin of the kidney stone can be made. The stone was probably formed due to a urinary tract infection, which was likely caused by a fracture of the ischiopubic ramus, which in turn can lead to urethral injuries. This trauma and its subsequent sequelae involved a lengthy process; in the case of kidney stones of infectious origin, this can be estimated at 4–6 weeks [69]. In more severe cases, as may be the case here, it could have resulted in renal failure.

The osteological study of the skeletal remains of the female buried in tomb AY-68 does not indicate a connection between health condition and the formation of the kidney stone. However, the study of *calculi* allowed the identification of an infectious process that would have otherwise remained undetected.

Infectious diseases have entailed a major challenge to human health throughout history. In 2019, 13.7 million global deaths were attributed to infections, and this figure was reported to have increased to 22% in 2021 due to COVID-19 [70]. Despite the impact of infections on morbidity and mortality, only 10% of the most common infections leave traces on the skeleton. In paleopathology this phenomenon is called “the iceberg principle” [71]. This leads to a significant limitation in assessing the health status of past populations. Urinary tract infections (UTIs) are an example of infectious diseases that remain “below the surface”. They are also highly prevalent, particularly in older female populations [72]. In a 2019 study of global mortality associated with 33 bacterial pathogens, infections by bacteria that affected the urinary tract system caused 4.9 deaths per 100,000 people [70]. The analysis of renal *calculi* provides indirect evidence of urinary tract infections, a condition that, thus so far, has not been addressed in prehistoric contexts.

The infections resulting in the *calculi* affected seriously the daily life of both individuals: fever, colic and, eventually, systemic infection leading to death. Most probably, their health issues required social care mechanisms addressed both to alleviate the symptoms and to compensate their contributions in terms of labor and other collective requirements. The attention and care given to the male buried in AY-60 to heal his wounds was particularly intensive and meaningful, as the long list of healed injuries was probably related with episodes of interpersonal physical violence required by the maintenance of his privileged position. This elite individual received persistent care despite his status of non-laborer [65, 73]. The female buried in AY-68 was an active laborer that also benefited from the care practices at work in Argaric society. Both examples give a previously unknown glimpse of what disease meant in terms of caregiving and caretaking in this prehistoric society.

## Data Availability

All relevant data are within the manuscript and its Supporting Information files.

## Acknowledgements

F.T. wishes to acknowledge the Vrije Universiteit Brussel for support, among others, through a Strategic Research Program awarded to his group.

The archaeological dimensions of this work were supported by the projects “Transformations: the formation of El Argar society (2200-2000 cal BCE) and the making of a symbolic and political order” (Spanish Ministry of Science, Innovation, and Universities, PID2023-146504NB-I00), *Grup de Recerca en Arqueoecologia Social Mediterrània* (Catalan Ministry of Research and Universities, AGAUR, 2021SGR0525), ICREA Acadèmia Programme (2024ICREA0082, R.M.) and Palarq Foundation (National Prize of Archaeology and Palaeontology). We would like to thank Miguel Valério, a member of the Almoloya-Bastida Project, for revising the English draft.

## Notes

### Competing Interest Statement

The authors have declared no competing interest.

### Funding Statement

Yes

### Author Declarations

This research did not involve living human participants and therefore did not require Institutional Review Board (IRB) approval. The study was conducted on archaeological human remains recovered during authorized excavations at La Almoloya (Murcia, Spain). All samples were analyzed with permission from the competent archaeological authorities and curating institutions, following all relevant national and regional regulations governing the study of human remains. No modern human data were collected.

